# Epidemiological methods provide target metrics and control parameters for multi-actor violent conflicts

**DOI:** 10.64898/2026.08.05.26359787

**Authors:** Michael Lazarus Smah, Kristian Skrede Gleditsch, Niall James MacKay

## Abstract

Violent conflicts increasingly involve multiple armed actors competing for influence over shared civilian populations, creating complex dynamics that challenge conventional security analysis and policy design. We present a framework that adapts epidemiological methods to model multi-actor violent conflict as an epidemic process, informed by the conflict landscape in Nigeria. We derive a basic insecurity reproduction number (*R*_0_), identify violence-free and persistent-violence equilibria, and introduce a novel Civilian Harm Index (CHI) to quantify humanitarian impact. Sensitivity analyses identify recruitment, ideological support from civilian populations, and abduction as the key drivers of conflict persistence and civilian harm. The framework reveals several counterintuitive findings. Interventions that most effectively suppress violence transmission are not necessarily those that minimise civilian harm, demonstrating that epidemic control and humanitarian protection may require distinct optimisation criteria. Likewise, interventions effective against one armed actor may be ineffective, or even counterproductive, when applied uniformly across groups. In addition, prisoner exchange and ransoming increase the persistence of violence and civilian harm. Although developed as an illustrative rather than predictive framework, our results show that epidemiological methods can provide quantitative metrics for evaluating intervention priorities and trade-offs in complex multi-actor conflicts.

## 1 Introduction

### 1.1 Background

Violent conflict is a persistent threat to human development and livelihoods. Armed conflicts inflict misery directly through loss of life and indirectly through severe social and economic consequences. Contemporary conflicts are often characterised by multiple armed actors, and multi-actor conflicts tend to be more intense, harder to resolve, and last longer^1–3^.

Conflicts involving actors with distinct ideological, political, or religious/ethnic objectives are also likely to be more difficult to resolve, since outcomes acceptable to some are likely to be rejected by others, and apparent gains or concessions from a government for one actor may encourage escalation by others.

Nigeria provides an important example of multi-actor conflict, highlighting common patterns in multi-actor conflicts elsewhere. Over the past 25 years, actors with different operational strategies and recruitment mechanisms have contributed to insecurities, including the jihadist insurgencies of Jama’atu Ahlis Sunna Lidda’awati wal-Jihad (JAS, popularly known as Boko Haram) and the Islamic State West Africa Province (ISWAP), armed bandit groups in the north-west, violent farmer–herder conflicts in the Middle Belt, separatist armed movements in the south-east, organised kidnapping networks, and community-based vigilante organisations^4–6^. These conflicts have profound humanitarian consequences, encompassing attacks, abductions and extortion, forced displacement and destruction of infrastructure, affecting millions. The United Nations Development Programme (UNDP) estimates that Boko Haram alone has caused tens of thousands of deaths and displaced millions across the Lake Chad Basin^6^. Similarly, kidnapping-for-ransom has evolved into a major source of insecurity in several Nigerian states, causing economic disruption while financially sustaining armed groups^7,8^. Many analyses of conflicts focus on individual groups in isolation. But even if groups may differ substantially in ideology, organisation and objectives, they are are often closely connected in strategic adaptation. They generally confront the same government and often compete over the same civilian populations for recruitment, coercion, intimidation, financial extraction and influence. The coexistence of multiple armed actors introduces complexity for policy and security responses, as such environments require governments and organisations to consider how actions and decisions affecting one armed actor may influence others and introduce potential unintended consequences. Governments thus face a dual challenge in allocating limited resources among different responses — i.e., military responses, policing, intelligence, hostage negotiations, de-radicalisation, and community and economic development — and in determining which interventions should be directed at which groups, and in what proportion^9,10^.

Traditional security approaches focusing on intelligence, military suppression and law enforcement have had mixed success, both in Nigeria and more generally^10^. Military responses alone rarely suffice to eliminate violence when recruitment channels remain open. Popular support – whether active and passive – from segments of the population is held to be key to effective insurgency in the theories of guerrilla warfare advanced by Mao and Che Guevara^11^. Sympathy, ideological alignment, ethnic affiliation, grievances, economic incentives and distrust of the state shape recruitment and operational resilience in north-eastern Nigeria^6,12^, in line with analyses of Afghanistan, Iraq, Colombia and elsewhere^9,10^. The limitations of traditional security approaches have motivated broader strategies of prevention, negotiation, rehabilitation and reintegration. Counter-radicalisation reduces ideological attraction through education, community engagement and economic empowerment^6^. Structured deradicalisation and rehabilitation programmes (e.g. the Nigerian Operation Safe Corridor (OPSC)^13^) seek to enable disengagement from extremism and promote re-integration, although critics have raised concerns regarding victim perceptions, accountability, and recidivism^14,15^. Negotiation plays an important role in conflict management, often reflected in prisoner exchanges, hostage negotiations, amnesty programmes and negotiated releases^16–18^.

However, actions that may well reduce conflict and its humanitarian impact in the short term can often generate unintended consequences, for example, by increasing the strategic value of hostage-taking or enhancing the legitimacy and bargaining power of armed actors. Suppressing one actor militarily can increase grievances that help recruitment in the longer term. Ransom payments may save lives in the short term but also create financial incentives that sustain future kidnapping operations^19,20^. And successful action against one actor may help create opportunities for other competing groups to gain prominence. Thus, violent insecurity is not solely a military/enforcement problem but also politics involving a population-level social process, underscoring how the relevance of von Clausewitz’s maxim that “war is a continuation of politics/policy by other means”^21^ extends to insurgencies.

### 1.2 Epidemic models for multi-actor conflict

Epstein^22^ highlights how quantitative models need not be “digital twins” but, provided they capture the core dynamics, can still provide a systematic framework for exploring interventions, identifying thresholds, evaluating trade-offs and informing evidence-based decision making in environments where intuition alone is insufficient or misleading.

The idea of applying epidemiological concepts to conflict is not new. The physicist and early pioneer of war studies Lewis Fry Richardson considered support for war akin to an epidemic^23^, and the similarity of the core dynamics has been explored recently^24^. Dynamical models have been advocated for various aspects of civil war and insurgency (for example^25,26^), but there has been surprisingly little use of established epidemiological methods^27,28^ in studying conflict^29,30^. Most existing mathematical models focus on a single armed actor or a homogeneous conflict environment and do not begin to capture the complexity of multi-actor conflicts.

We provide as a “proof of concept” the use of dynamical epidemiological modelling as a tool for analysing multi-actor conflict. We develop a mathematical framework, in the form of a multi-compartment dynamical system, for analysing violent conflict as an epidemic-type process. Armed groups are represented as interacting actors that recruit, coerce, radicalise, abduct, and compete for influence over a homogeneous civilian population. The framework integrates violent and non-violent intervention mechanisms, including military suppression, arrests, deradicalisation, reintegration, prisoner exchange, hostage negotiation, and ransom-related processes. We can observe standard epidemiological phenomena: an initial epidemic controlled by a reproduction number, leading to an endemic state of persistent violence. The basic epidemiological approach is to control the initial epidemic through the reproduction number, and we analyse this, identifying threshold conditions governing the persistence and control of the conflict. Then, to enable action based on the model, we create a novel Civilian Harm Index (CHI), a measure of the instantaneous and cumulative harm being done to the population by the conflict. This enables us to analyse the human value of various possible interventions, their relative efficacy, and the trade-offs between them.

## 2 Model Formulation

We model violent conflict as an epidemic-style dynamical system involving two distinct armed actor groups that interact indirectly through competition for a shared civilian population. Epidemic models typically use a set of compartments appropriate to the progression of the disease^31^ (for example, susceptible-exposed-infectious-removed), and we create here a set of compartments inspired by the operational chharacteristics of groups in the Nigerian conflict, but which can easily be adapted to others:

- **Group 1 (G1):** G1 (predatory recruiters – ideologically-based) combines violent attacks with coercive and ideological recruitment. The group can both murder and abduct civilians. Abducted individuals enter the abducted compartment *A*_1_, where they may subsequently be indoctrinated and recruited into the active armed population *I*_1_, or killed while in captivity. G1 does not engage in ransom negotiations or financial exchange for captives, and primarily sustains itself through forced recruitment and ideological attraction from sympathetic populations.
- **Group 2 (G2):** G2 (mixed criminal/ransom – financially-based) combines violent activity with financially-motivated criminal operations. Similar to G1, it can both murder and abduct civilians. However, G2 also actively engages in ransom negotiations and prisoner exchange, allowing captives to generate financial or strategic value. Abducted individuals enter the exposed compartment *A*_2_, where they may be released through ransom-based negotiation or killed in captivity. Recruitment into G2 is driven by financial vulnerability, economic incentives, and criminal opportunism.

G1 is typical of the mode of operation of Boko Haram and other jihadist factions in north-eastern Nigeria. They employed armed raids on communities, with mass abductions and use of abductees for forced labour, forced marriage, and coerced religious conversion^32^. Some, particularly children and young people, have been forcibly recruited into the group and trained or coerced to participate in attacks^33^. Captives have also been used as bargaining for prisoner exchanges with the Nigerian government^34^, and although ransom-taking has not been widely considered a motivation for Boko Haram activities, there is evidence of ransom-payment and exchange of captives^35^. By contrast, G2 is typical of armed bandit groups operating mainly in the Middle Belt and north-western Nigeria. They are highly fragmented and are generally motivated more by economic gain through kidnapping for ransom rather than by ideological objectives^36^.

The interaction between the two armed groups is indirect and occurs through competition for the susceptible population. This is analogous to co-circulating epidemic systems in which multiple infectious agents compete for susceptible hosts without direct interaction between strains.

The model, including all its compartments and parameters governing flow between them, is represented by the schematic flow diagram in Figure 1. To ensure bounded transmission dynamics and numerical stability, we adopt a frequency-dependent interaction structure in which recruitment, radicalisation, and abduction processes are normalised by the total population size, N(t). The full equations of the dynamical system, along with the full list of model parameters and their epidemiological/security interpretations, associated units and baseline values given in the Supplementary Materials.

**Figure 1.**
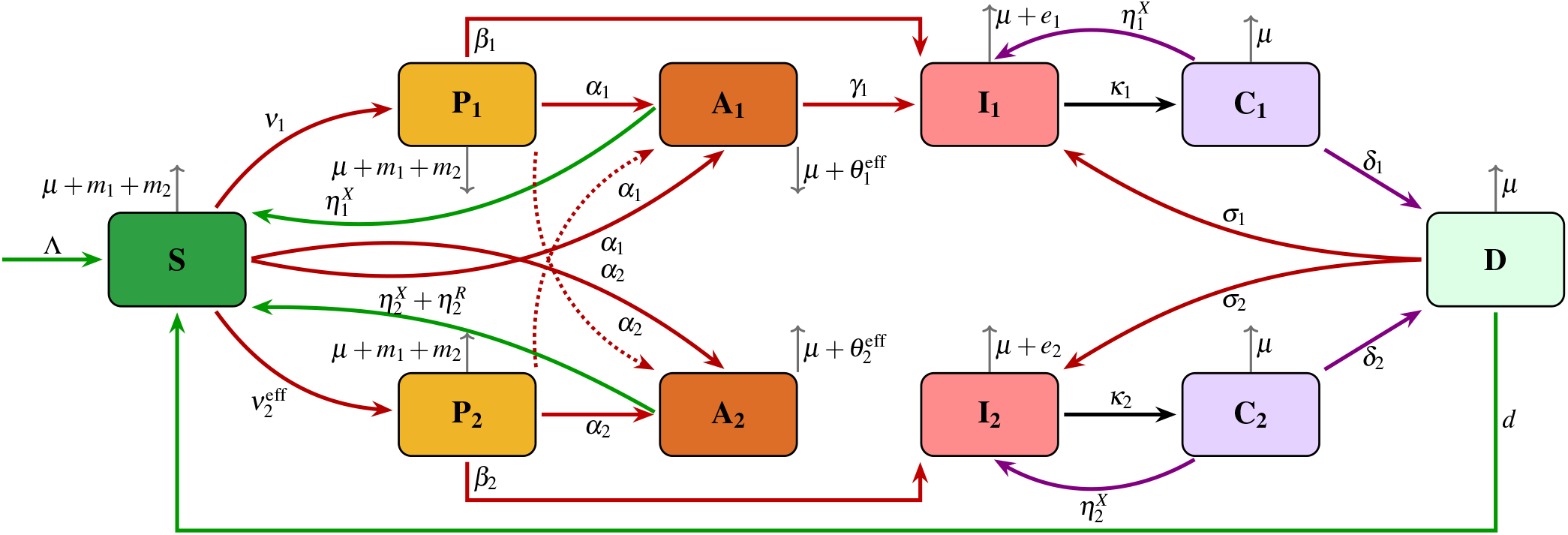
Compartmental flow diagram of the two-group violent insecurity dynamics model. The direction of the arrows indicates transitions between states.

The simulation outcomes are intended as illustrative demonstrations of the modelling framework rather than quantitative forecasts of a specific conflict.

### 2.1 Key Assumptions

The population is divided into

- *S*(*t*): Susceptible civilians (vulnerable to forced recruitment or violence. and without active sympathies).
- *P*_1_(*t*), *P*_2_(*t*): Populations sympathetic to armed groups 1 and 2 respectively.
- *A*_1_(*t*), *A*_2_(*t*): People abducted and held captive by G1, G2.
- *I*_1_(*t*), *I*_2_(*t*): Active violent members of groups 1, 2.
- *C*_1_(*t*), *C*_2_(*t*): Active members arrested/captured.
- *D*(*t*): Deradicalised individuals.

New recruitment is directly from sympathisers *P*_1_(*t*), *P*_2_(*t*) while G1 also recruits from abducted *A*_1_(*t*). All variables represent non-negative population counts at time *t*, large enough to be modelled as continuous, and the total population is

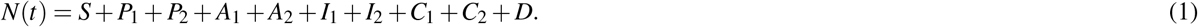

1. Civilians *S*, with birth rate Λ, are unsympathetic towards armed groups.
2. All individuals in *S, P*_1_, and *P*_2_ are vulnerable to murder *m*_*g*_ and abduction *α*_*g*_ by all armed groups (*g* = 1, 2).
3. Individuals in *S* become sympathetic towards group *g* at rate *ν*_*g*_.
4. Voluntary recruitment into group *g* occurs from individuals sympathetic towards group *g* (*P*_*g*_) at rate *β*_*g*_.
5. Recruitment into armed group *I*_1_ also occurs by radicalisation *γ*_1_ of abducted individuals in (*A*_1_).
6. Captives *C*_*g*_ of either group may be released through prisoner exchange 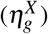 or undergo deradicalisation at rate *δ*_*g*_.
7. Deradicalised individuals may successfully reintegrate (*d*) into *S*, while others may relapse *σ*_*g*_ into active violence.
8. Unsuccessfully deradicalised individuals may rejoin any of the available armed groups.
9. All populations experience natural mortality at rate *µ*; additional mortality arises from violent attacks (*m*_*g*_), in captivity (*θ*_*g*_), and as targeted elimination *e*_*g*_ of active militants.
10. The terms *m*_*g*_ represent direct civilian mortality during attacks and therefore only apply to non-combatant compartments.
11. Negotiation and prisoner exchange reduce mortality in captivity.

To account for the effect of negotiation and prisoner exchange on captivity-related mortality, we define effective killing rates in captivity as

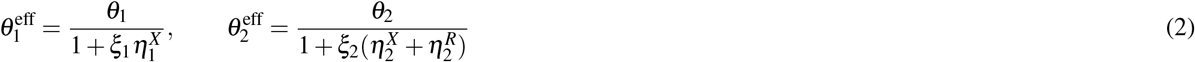

where *ξ*_*g*_ *>* 0 measures the effectiveness of negotiation and exchange mechanisms in reducing killings of captives by group *g*.

To capture the economic attraction of successful ransom operations, we define an effective voluntary sympathy 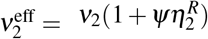, where *Ψ >* 0 measures how successful ransom activity amplifies interest for the financially-motivated armed actor group.

### 2.2 Model Analysis

The model was analysed using a combination of equilibrium analysis, threshold analysis, numerical simulation, uncertainty quantification, and intervention evaluation (Supplementary Materials).

Two equilibrium states were obtained as fixed points (constant solutions) of the dynamical system. The violence-free equilibrium (VFE) has no active violence, although susceptible and sympathiser populations exist. The persistent violence equilibrium (PVE) has non-trivial numbers in all compartments and indicates sustained endemic violence. The VFE and PVE were used as reference states for threshold, stability, and intervention analyses.

First, the basic (‘insecurity’) reproduction number (*R*_0_), which controls epidemic (exponentially growing) violence from the VFE was derived using the next-generation matrix framework^37^. The model equations were partitioned into recruitment and transition components, and the corresponding Jacobians evaluated at the VFE to construct the reproduction number.

To evaluate societal consequences beyond transmission dynamics, we introduced a Civilian Harm Index (CHI), an instantaneous rate per head of civilian population which combines multiple dimensions of civilian burden, including mortality, abduction, forced radicalisation, and ransom costs (see supplementary). Instantaneous civilian harm was also integrated over time to obtain a cumulative CHI.

Parameter uncertainty was explored using Latin Hypercube Sampling, and parameter influence was quantified using complementary rank-based and variance-based sensitivity measures. Intervention scenarios were implemented through systematic modification of policy-relevant parameters representing prevention, enforcement, negotiation, rehabilitation and suppression mechanisms, and performance was evaluated using cumulative CHI.

Prevention of an epidemic of violence requires reduction of the reproduction number to *R*_0_ < 1, which may be unachievable through single interventions. We therefore performed a systematic single-parameter threshold exploration. Each controllable parameter was varied independently while all remaining parameters were fixed at baseline values. Parameters representing recruitment, sympathy generation, contact and relapse following deradicalisation were reduced to their theoretical lower bound of zero, whereas control-related parameters (capture, elimination, deradicalisation, reintegration, negotiation, and captivity exit) were increased from baseline to their upper bound of one. For each intervention, *R*_0_ was recomputed and the minimum achievable value recorded. The objective was to determine whether any individual intervention could independently force *R*_0_ < 1.

For numerical simulation, the initial population state was specified to be *S*(0) = 500000, *P*_1_(0) = 20000, *P*_2_(0) = 15000, *I*_1_(0) = 50, *I*_2_(0) = 40 with *A*_1_(0) = *A*_2_(0) − *C*_1_(0) = *C*_2_(0) = *D*(0)) = 0, representing a predominantly susceptible population with existing ideological support and a small initial active insecurity presence.

Numerical simulations were performed in `R` using the `deSolve` package to run a system of ordinary differential equations. The dynamical system of ODEs was solved using the adaptive `lsoda` algorithm, which automatically switches between stiff and non-stiff integration methods depending on the local characteristics of the system. Numerical integration was conducted over the interval *t ∈* [0, 1500], using a step increment of 0.2 time units and solver tolerances `rtol` = 10^*−*8^, `atol` = 10^*−*10^, with a maximum of 500000 integration steps to ensure numerical stability and convergence.

## 3 Results

Under the baseline parameter configuration, *R*_0_ = 2.496, leading to persistent insecurity and enabling meaningful investigation of long-term humanitarian harm, sensitivity behaviour, and intervention performance.

Figure 2 presents the decomposition of this baseline *R*_0_ into its constituent next-generation components. The dominant contribution arises from the within-group reproduction pathways arising from the groups’ sympathisers. G1 exhibited the largest self-sustaining contribution (*R*_11_ = 2.487), including voluntary recruitment from sympathisers and forced recruitment from abducted individuals. This is followed by G2 recruitment from its sympathisers (*R*_22_ = 1.752). The cross-group coupling terms generated through relapse from unsuccessful rehabilitation are comparatively small, with recruitment to G1 from original G2 members (*R*_12_ = 0.116), and for G2 from G1 (*R*_21_ = 0.057).

**Figure 2.**
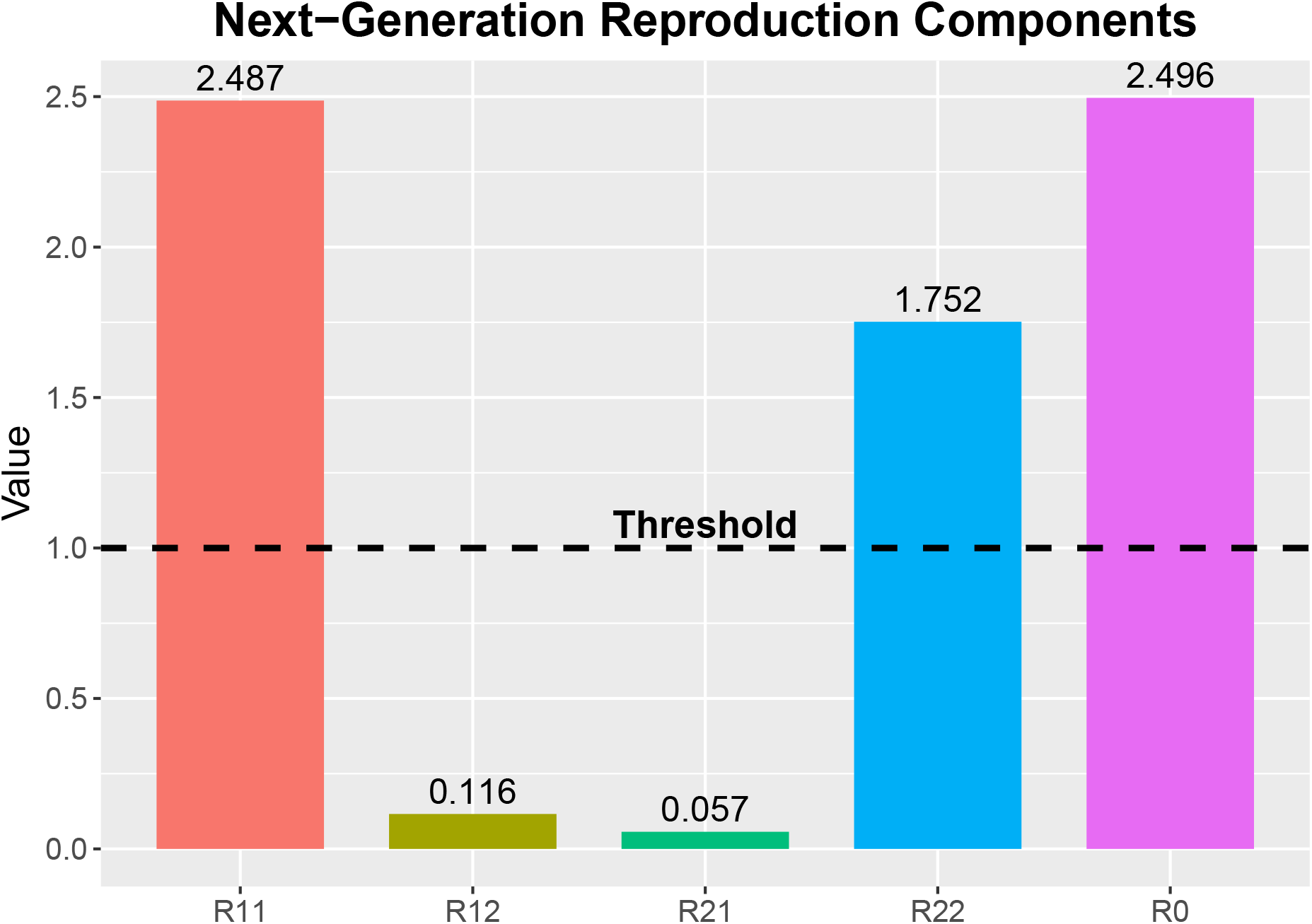
Decomposition of reproduction components and overall reproduction threshold. Contribution of within-group and cross-group pathways to the coupled basic (’insecurity’) reproduction number (*R*_0_). Bars show the next-generation matrix components (*R*_11_), (*R*_12_), (*R*_21_), and (*R*_22_) and the resulting overall *R*_0_. The dashed line denotes the threshold (*R*_0_ = 1). *R*_11_ and *R*_22_ represent within-group reproduction, while *R*_12_ and *R*_21_ capture cross-group feedback through rehabilitation and relapse.

Table 1 shows the results of the systematic single-parameter threshold exploration. No single intervention reduced *R*_0_ below one, so that threshold reduction requires coordinated intervention across several mechanisms simultaneously.

**Table 1.**
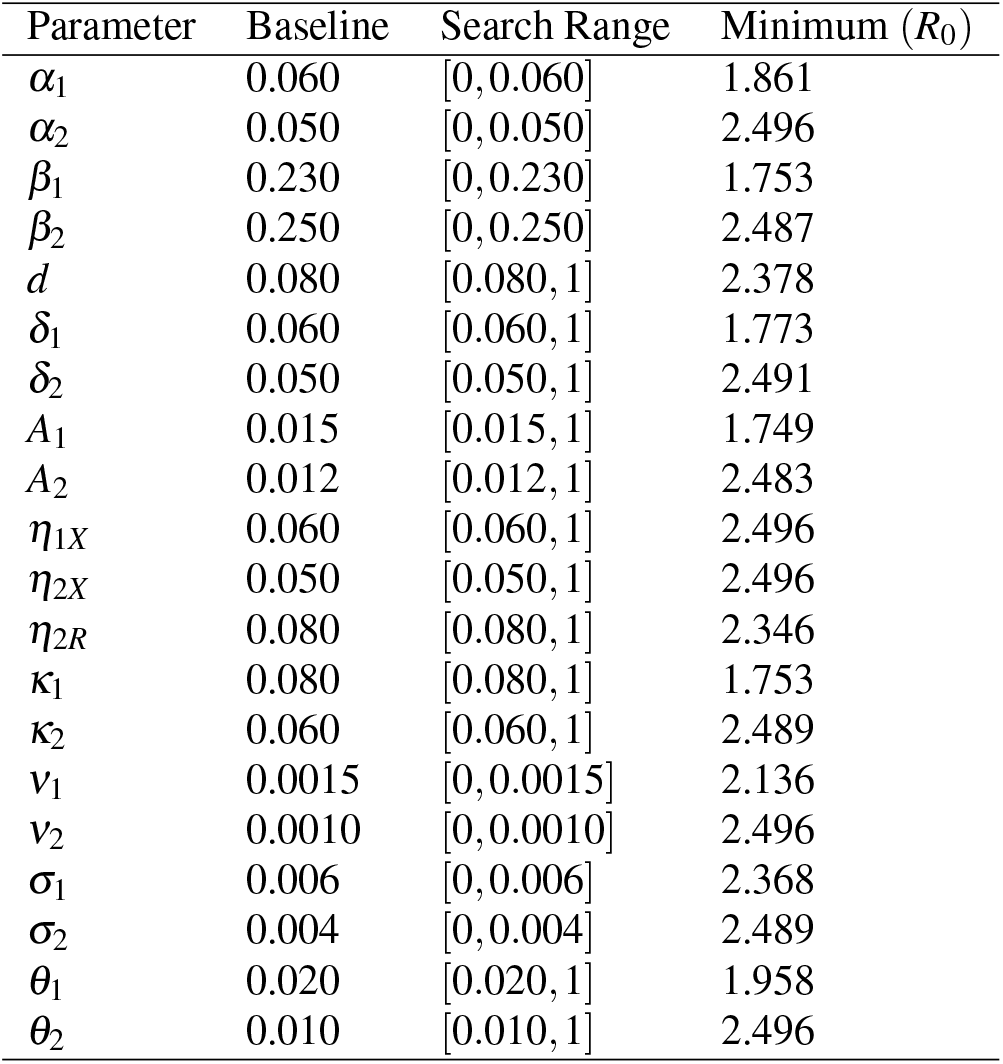
Single-parameter intervention screening for threshold reduction. Each parameter was varied independently while all other parameters remained fixed at baseline values. Reduction parameters were explored from baseline to (0), while enhancement parameters were explored from baseline to (1).

Figure 3 illustrates the baseline model dynamics (left), and the dynamics of abducted and active members of Groups 1 and 2 (right). The initial expansion phase is characterised by growth in sympathisers with corresponding fall in susceptibles, followed by delayed increases in active violent actors. However, the susceptible population partially recovers and approaches a stable positive level, indicating sustained but non-collapsing insecurity dynamics. This suggests persistent violence coexisting with long-term demographic stability. The abducted and active violent subpopulations emerged after a short delay and increased once sufficient passive support accumulated. G1 generated an earlier and stronger active wave, while G2 displayed slower but sustained growth. The population of G1 sympathisers rapidly increases, but falls below the pool of G2 sympathisers in the long run. The high population of G2 sympathisers, despite the lower baseline rate compared to G1, is linked to the effects of ransom payments, attracting more recruits. Persistence of violent insecurity is maintained through feedback among recruitment, captivity, rehabilitation, and relapse processes rather than direct transmission alone.

**Figure 3.**
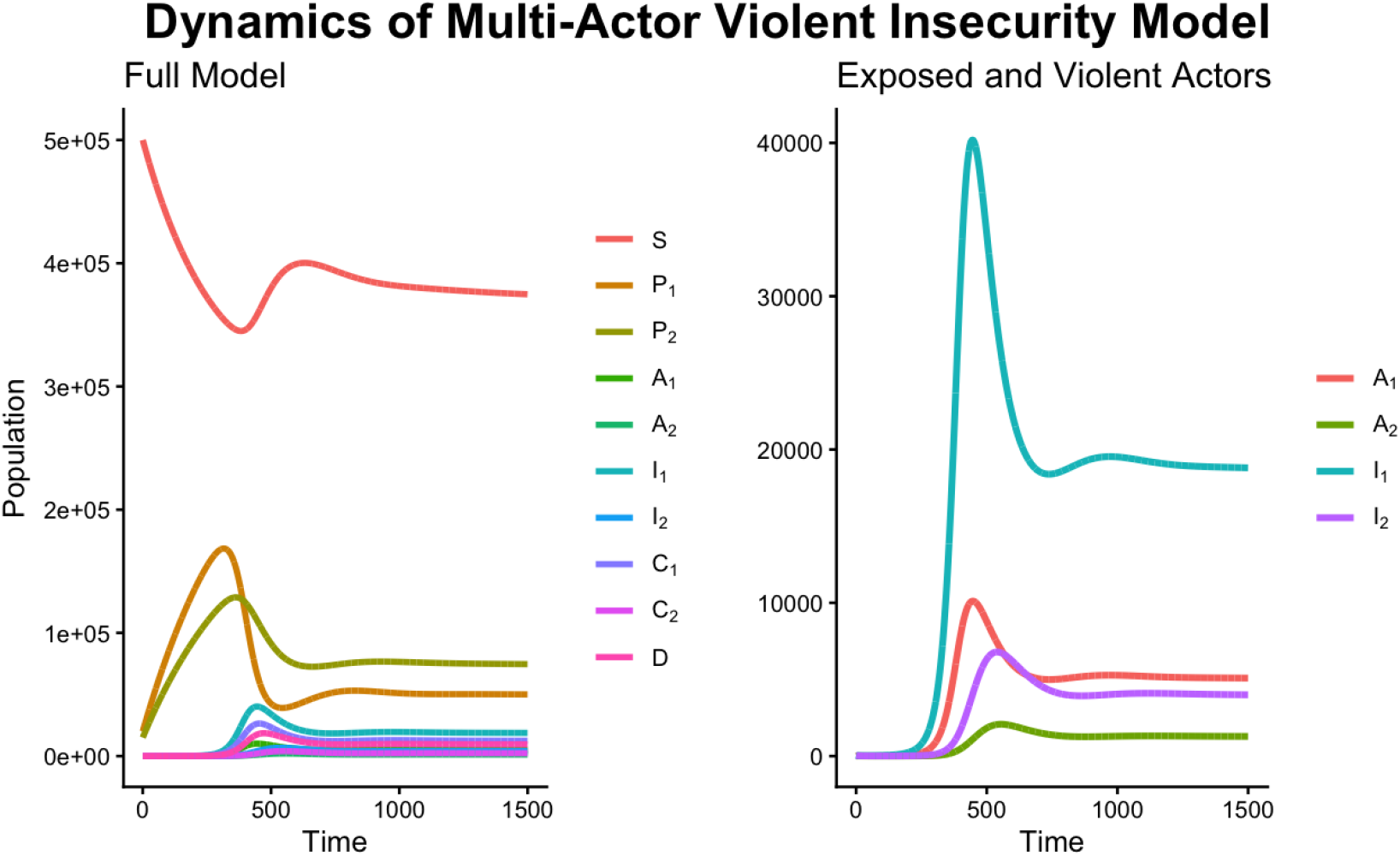
Baseline dynamics of the violent insecurity model showing the evolution of population compartments over the simulation time.

Global sensitivity analysis was performed using Partial Rank Correlation Coefficients (PRCC) to identify parameters with the greatest influence on both *R*_0_ and the cumulative harm. Figure 4 presents the resulting parameter rankings.

**Figure 4.**
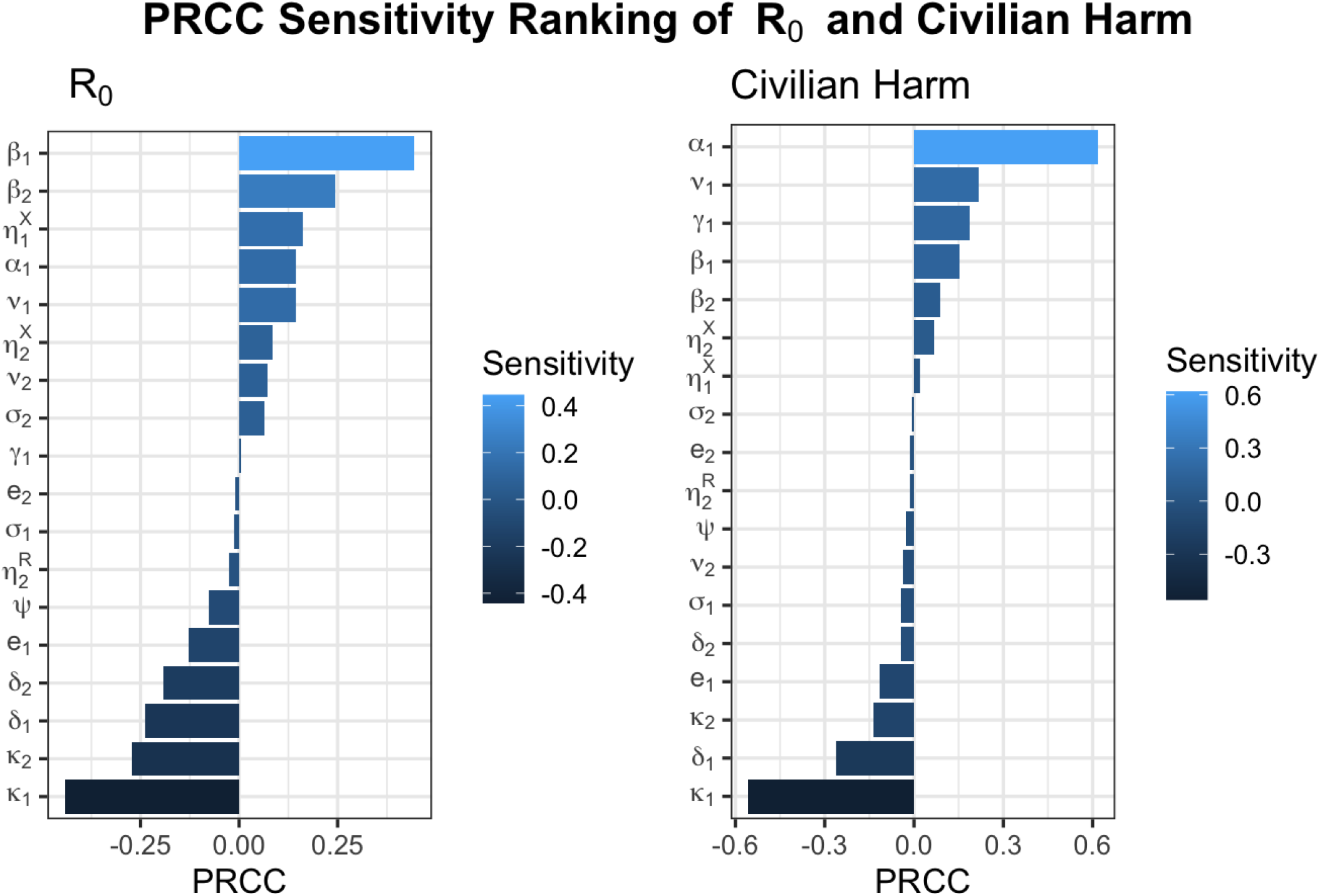
Spearman PRCC sensitivity ranking for the basic (’insecurity’) reproduction number (left) and cumulative Civilian Harm Index (right). The lengths of the bars correspond to the effects of the parameters on the entire quantity. Positive (resp., negative) coefficients mean that increasing the parameter would increase (resp. decrease) the quantity (*R*_0_ or cumulative CHI).

The top five parameters with the strongest influence on *R*_0_ are voluntary recruitment into G1 (*β*_1_), into G2 (*β*_2_), negotiated prisoner exchange with G1 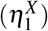 abduction by G1 (*α*_1_), and generation of sympathisers for G1 (*ν*_1_). Increasing any of these increases *R*_0_ and favours the persistence and spread of insecurity. In contrast the parameters with the strongest negative influence on *R*_0_ are arrests of active G1 (*k*_1_) and G2 members (*k*_2_), de-radicalisation of arrested G1 (*δ*_1_) and G2 members (*δ*_2_), and military elimination of active G1 members (*e*_1_). Increasing any of these reduces *R*_0_.

Sensitivity analysis of the cumulative CHI reveals a different hierarchy. The dominant contributor is abduction by G1 (*α*_1_), followed by generation of sympathisers for G1 (*ν*_1_), radicalisation of abducted individuals into G1 (*γ*_1_), and voluntary recruitment into G1 (*β*_1_) and G2 (*β*_2_). Thus abduction, indoctrination of civilians into supporting the ideology of G1, and subsequent voluntary enlistment into the active group are the principal drivers of cumulative harm. The strongest reductions in civilian harm (in the order of the top five parameters) are due to arrests of active G1 members (*k*_1_), de-radicalisation of arrested G1 members (*δ*_1_), arrests of active G2 members (*k*_2_), military elimination of G1, and de-radicalisation of arrested G2 members (*δ*_2_). These apply both cumulatively and instantaneously when in the PVE.

Importantly, the sensitivity rankings for *R*_0_ and cumulative CHI are not identical. Parameters that most effectively suppress insecurity transmission are not necessarily those that minimise civilian harm. This distinction highlights the need for intervention strategies that jointly optimise transmission control and humanitarian protection.

We now compare the effects of various interventions on the CHI. Figure 5 shows both intervention trajectories and percentage reduction relative to baseline. Among all interventions, increasing arrest in both groups simultaneously shows the highest reduction in cumulative civilian harm, achieving a little above 60% reduction relative to the baseline. Reducing ideological sympathy generated the second highest reduction, followed by increased military pressure, and de-radicalisation in that order. Increasing ransom payment and prisoner exchange show negative impacts, with the highest negative impact from prisoner exchange relative to the baseline.

**Figure 5.**
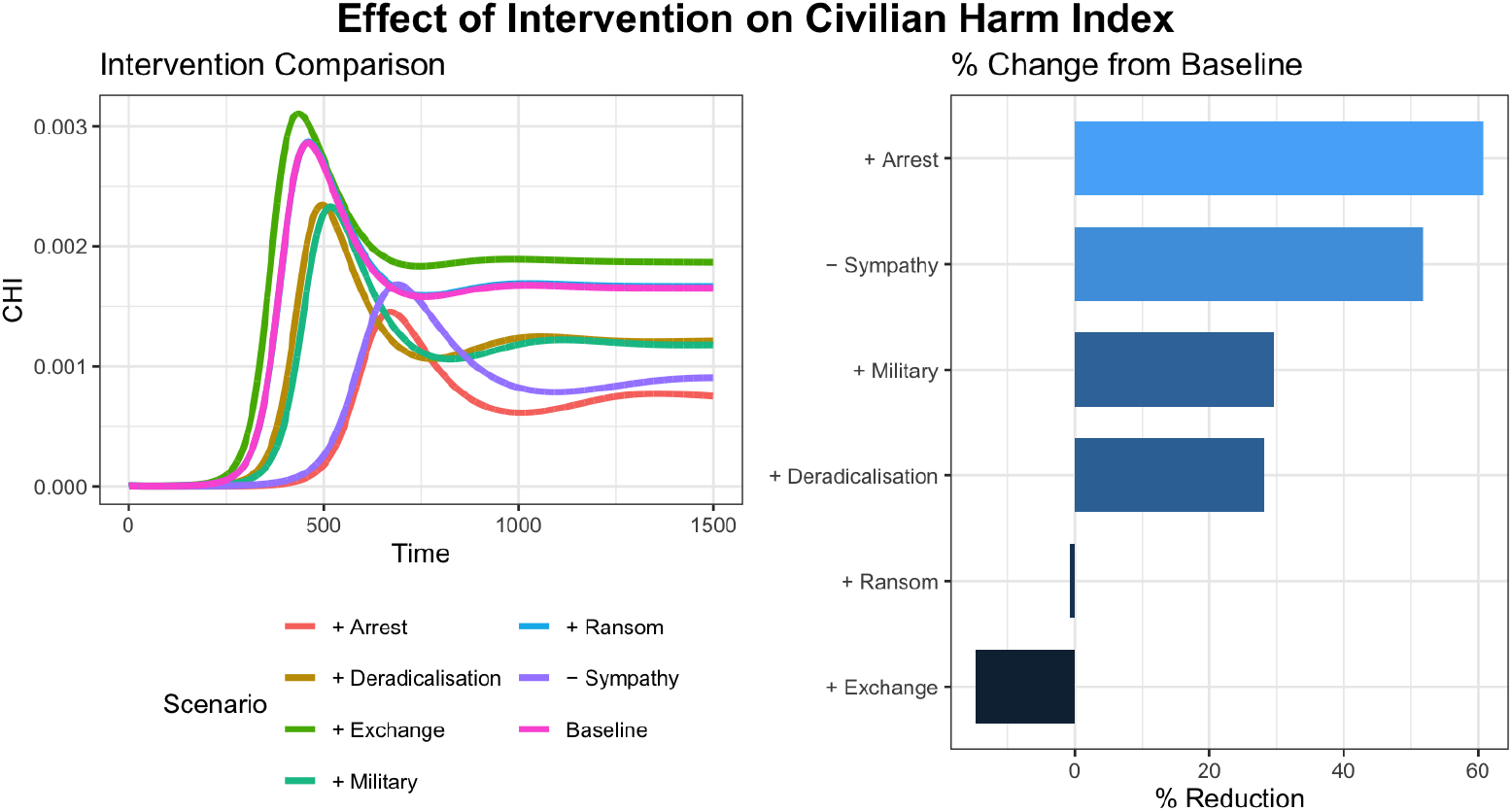
Intervention comparison based on the Civilian Harm Index over time. The left panel shows the intervention trajectories; the right panel shows the percentage reduction relative to baseline. Plus (+) sign indicates increase in the corresponding parameter, while minus (−) sign mean reduction in the parameter. All changes in the parameters were ± the baseline values.

The resulting persistent violence equilibrium (PVE) is 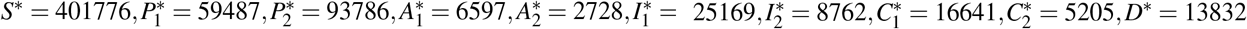 indicating coexistence of susceptible individuals, ideological support populations, abducted individuals, active violent actors, captured, and rehabilitated individuals under sustained insecurity conditions. In particular, both active members’ compartments remained positive 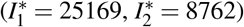, demonstrating that the system converges to a PVE rather than returning to the violence-free VFE.

Local stability of the persistence equilibrium was investigated by evaluating the Jacobian matrix at the PVE and computing its eigenvalues numerically. All eigenvalues have strictly negative real parts, with dominant eigenvalue *λ*_max_ = *−*0.00135, so that the PVE is locally stable. Four eigenvalues were complex, indicating the inspiral (damped oscillatory) behaviour evident in Figs. 3 & 5.

## 4 Discussion

We have demonstrated that standard epidemiological methods provide a productive modelling framework for violent multi-actor conflict. Our results offer a plausible simulation, and jointly indicate that persistent insecurity is governed primarily by recruitment, ideological support, captivity, and disengagement mechanisms.

The existence of the PVE implies that under the baseline parameter values, insecurity becomes self-sustaining. Temporary reductions in recruitment, violence, or active participation are insufficient to permanently disrupt long-run insecurity dynamics unless interventions alter the underlying mechanisms sufficiently to move the system outside the PVE’s basin of attraction by reducing *R*_0_ below one. Transient improvement might occur due to the oscillatory behaviour associated with the complex eigenvalues, but this should not be mistaken for solution of the problem. Further, the relatively small magnitude of the dominant eigenvalue implies slow convergence, with the PVE resilient to short-term disturbances. Sustained interventions may be necessary to achieve meaningful long-term reduction in insecurity burden.

### 4.1 Policy Implications

The threshold exploration results have important implications for the design of insecurity-mitigation strategies in multi-actor environments. Unlike classical single-group transmission systems, our model demonstrates that reducing the reproduction potential of one actor alone may not be sufficient to eliminate persistent violence.

Responses to conflict should move beyond actor-specific approaches and instead adopt system-level intervention frameworks. In practical terms, strategies focused exclusively on suppressing one organisation through military pressure, targeted elimination, arrests, or negotiation may generate limited long-term impact if competing or complementary actors continue to maintain recruitment and operational capacity.

The inability of any single parameter to reduce *R*_0_ below unity further indicates that persistence of violence emerges from the combined influence of multiple mechanisms rather than a dominant driver. Thus, the elimination of insecurity requires coordinated intervention across the entire violence ecosystem, including reducing sympathy generation, disrupting recruitment channels, strengthening de-radicalisation programmes, improving reintegration pathways, limiting relapse, and simultaneously increasing operational disruption across all active groups.

This supports an integrated security policy in which military operations, intelligence activities, community engagement, rehabilitation programmes, economic interventions, and hostage-response mechanisms are implemented jointly rather than independently. Such coordinated approaches are more likely to shift the system below the persistence threshold and prevent replacement effects whereby suppression of one actor unintentionally creates opportunities for another. We note that this is consistent with existing arguments that effective counterinsurgency can often be strengthened by emulating strategies in United Nations integrated peacekeeping missions, beyond the narrow provision of security Friis01022010.

More broadly, we hope to have demonstrated that treating multi-actor violence as an epidemic and then an endemic disease provides a sound framework for the analysis of interventions. Our results suggest that analyses of insecurity in multi-actor populations should explicitly account for all active groups and their interactions to determine ‘which intervention is best for whom’ rather than focusing on any particular actor in isolation, or implementing intervention uniformly. The design of intervention strategies should take into account the relative effectiveness and thus the priority of parameter reductions across all active agents of insecurity. Models and policies that ignore cross-group dynamics may underestimate the resilience of insecurity and overestimate the effectiveness of single-target interventions.

The model parameters in this study have not been calibrated to empirical data, so the results should not be interpreted as quantitative predictions for any particular conflict. Rather, the model is intended as proof of concept for a broader policy-modelling framework. Its purpose is to examine how different mechanisms of recruitment, ideological support, violence, captivity, rehabilitation and relapse interact, and how interventions acting on these mechanisms can produce different system-level consequences.

The present model assumes constant demographics and conflict parameters, homogeneous mixing, and does not consider spatial heterogeneity, evolving alliances, political negotiations, or foreign intervention. However, it would be straightforward to adapt the model to the characteristics of a specific conflict. Compartment types and the dynamics of movement between them could be varied, for example, to accommodate ‘spoilers’^38^, in which settlement with one actor may be exploited by a challenger seeking to undermine peace agreements, recruit defectors and outcompete other challengers. Above all, future work should aim at realistic parameter calibration sufficient to enable some degree of predictive capability and its testing.

To finish, we reprise some potentially important policy principles suggested by our qualitative findings. First, interventions targeting a single actor or mechanism may be insufficient to defeat insecurity from the group when alternative pathways from another actor group sustain violence. Second, interventions that reduce transmission or persistence may not be those that minimise immediate civilian harm. Third, interventions affecting negotiation, ransom and captivity may involve trade-offs between immediate humanitarian protection and defeating longer-term incentives within the conflict system.

## Data Availability

This study focuses on the theoretical development of a mathematical model for multi-actor conflicts as a contagious spread of infectious diseases. No data was collected and analysed. The R code for the model simulation will be made accessible upon publication.

## Acknowledgements

M.L.S. is funded by the London Mathematical Society Early Career Fellowship.

## Author contributions statement

M.L.S. conceptualised the framework, generated the schematic diagram and the numerical analysis. M.L.S. & N.J.M. developed the model and performed the mathematical analysis. K.S.G. provided a broader context on conflict. M.L.S., N.J.M. & K.S.G. discussed the results and produced the final manuscript.

## Data availability

The R code for the model simulation will be made accessible upon publication.

## Ethics declarations

This study focuses on the theoretical development of a mathematical model for multi-actor conflicts. No data was collected and analysed.

