## Supplementary Information for "Epidemiological methods provide target metrics and control parameters for multi-actor violent conflicts"

### Supporting Information

#### 1 Model Parameters and Interpretation

The model parameters, their epidemiological/security interpretations, and associated units are summarised in Table 1.

**Table 1.** Model parameters and their interpretations.

| Parameter | Meaning | Units |
|---|---|---|
| $\Lambda$ | Birth and migration into the susceptible population | persons/time |
| $\mu$ | Natural mortality rate | 1/time |
| $\nu_1$ | Rate at which susceptibles become sympathisers of Group 1 | 1/time |
| $\nu_2$ | Rate at which susceptibles become sympathisers of Group 2 | 1/time |
| $\beta_1$ | Active recruitment rate from sympathisers $P_1$ into violent actors $I_1$ | 1/time |
| $\beta_2$ | Active recruitment rate from sympathisers $P_2$ into violent actors $I_2$ | 1/time |
| $\alpha_1$ | Abduction/contact rate associated with Group 1 | 1/time |
| $\alpha_2$ | Abduction/contact rate associated with Group 2 | 1/time |
| $\gamma_1$ | Radicalisation rate of abducted individuals into Group 1 | 1/time |
| $\eta_1^X$ | Prisoner exchange/release rate for Group 1 | 1/time |
| $\eta_2^X$ | Prisoner exchange/release rate for Group 2 | 1/time |
| $\eta_2^R$ | Negotiated ransom-release rate for Group 2 | 1/time |
| $\xi_1$ | Negotiation effectiveness coefficient for Group 1 | dimensionless |
| $\xi_2$ | Negotiation effectiveness coefficient for Group 2 | dimensionless |
| $\theta_1$ | Baseline captivity mortality rate among abductees of Group 1 | 1/time |
| $\theta_2$ | Baseline captivity mortality rate among abductees of Group 2 | 1/time |
| $\kappa_1$ | Capture/arrest rate of active Group 1 members | 1/time |
| $\kappa_2$ | Capture/arrest rate of active Group 2 members | 1/time |
| $e_1$ | Elimination rate of active Group 1 members | 1/time |
| $e_2$ | Elimination rate of active Group 2 members | 1/time |
| $\delta_1$ | De-radicalisation rate of captured Group 1 members | 1/time |
| $\delta_2$ | De-radicalisation rate of captured Group 2 members | 1/time |
| $d$ | Reintegration/recovery rate of de-radicalised individuals | 1/time |
| $\sigma_1$ | Relapse rate of de-radicalised individuals into Group 1 | 1/time |
| $\sigma_2$ | Relapse rate of de-radicalised individuals into Group 2 | 1/time |
| $\psi$ | Recruitment amplification effect induced by ransom success | dimensionless |

The population is divided into the following compartments:

- $S(t)$: Susceptible civilians (individuals vulnerable to recruitment or violence).
- $P_1(t)$, $P_2(t)$: Sympathetic populations to armed Group 1 and Group 2, respectively.
- $A_1(t)$, $A_2(t)$: Abducted/exposed individuals held by Group 1 and Group 2.
- $I_1(t)$, $I_2(t)$: Active violent members of Group 1 and Group 2.
- $C_1(t)$, $C_2(t)$: Arrested/captured members of Group 1 and Group 2.
- New recruitment into Group 1 are directly from the sympathisers, $P_1(t)$ and from the abducted $A_1(t)$.
- New recruitment into Group 2 are directly from the sympathisers, $P_2(t)$.
- $D(t)$: Deradicalised individuals.

All variables represent non-negative population counts at time $t$, and the total population is

$$N(t) = S + P_1 + P_2 + A_1 + A_2 + I_1 + I_2 + C_1 + C_2 + D. \tag{1}$$

This model is represented by the schematic flow diagram in Figure 1.

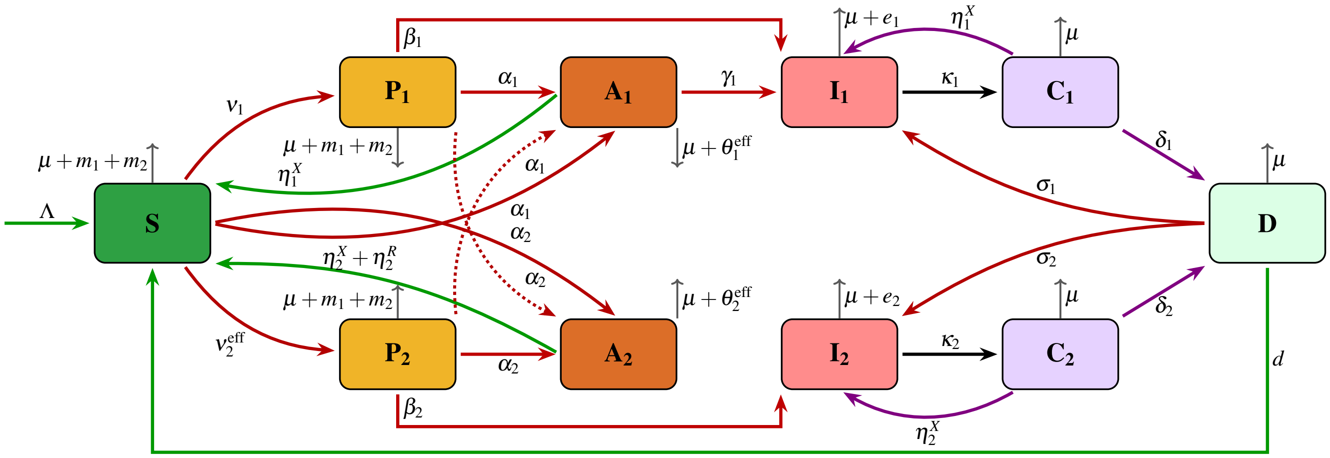


**Figure 1.** Compartmental flow diagram of the two-group violent insecurity dynamics model. The direction of the arrows indicates transitions between states.

###### 1.0.1 Governing Equations

To ensure bounded transmission dynamics and numerical stability, we adopt a frequency-dependent interaction structure in which recruitment, radicalisation, and abduction processes are normalised by the total population size, N(t).

The dynamics are governed by the system of ordinary differential equations (2 - 11).

$$\frac{dS}{dt} = \Lambda - \nu_1 S - \nu_2^{\text{eff}} S - (\alpha_1 + m_1)\frac{SI_1}{N} - (\alpha_2 + m_2)\frac{SI_2}{N} + \eta_1^X A_1 + (\eta_2^X + \eta_2^R) A_2 + dD - \mu S, \tag{2}$$

$$\frac{dP_1}{dt} = \nu_1 S - (\beta_1 + \alpha_1 + m_1)\frac{P_1 I_1}{N} - (\alpha_2 + m_2)\frac{P_1 I_2}{N} - \mu P_1, \tag{3}$$

$$\frac{dP_2}{dt} = \nu_2^{\text{eff}} S - (\beta_2 + \alpha_2 + m_2)\frac{P_2 I_2}{N} - (\alpha_1 + m_1)\frac{P_2 I_1}{N} - \mu P_2, \tag{4}$$

$$\frac{dA_1}{dt} = \frac{(S + P_1 + P_2)\alpha_1 I_1}{N} - (\gamma_1 + \eta_1^X + \theta_1^{\text{eff}} + \mu) A_1, \tag{5}$$

$$\frac{dA_2}{dt} = \frac{(S + P_1 + P_2)\alpha_2 I_2}{N} - (\eta_2^X + \eta_2^R + \theta_2^{\text{eff}} + \mu) A_2, \tag{6}$$

$$\frac{dI_1}{dt} = \frac{\beta_1 P_1 I_1}{N} + \gamma_1 A_1 + \eta_1^X C_1 + \sigma_1 D - (\kappa_1 + e_1 + \mu) I_1, \tag{7}$$

$$\frac{dI_2}{dt} = \frac{\beta_2 P_2 I_2}{N} + \eta_2^X C_2 + \sigma_2 D - (\kappa_2 + e_2 + \mu) I_2, \tag{8}$$

$$\frac{dC_1}{dt} = \kappa_1 I_1 - (\eta_1^X + \delta_1 + \mu) C_1, \tag{9}$$

$$\frac{dC_2}{dt} = \kappa_2 I_2 - (\eta_2^X + \delta_2 + \mu) C_2, \tag{10}$$

$$\frac{dD}{dt} = \delta_1 C_1 + \delta_2 C_2 - (d + \sigma_1 + \sigma_2 + \mu) D. \tag{11}$$

#### 2 Derivation of the basic ('insecurity') Reproduction Number

We define insecurity infection as *entry into the violent-actor life cycle*. An individual is considered infected once they become an active violent actor, undergo forced radicalisation under the operational control of a violent organisation, or enter a compartment that can subsequently contribute to the regeneration of future active violent actors. Under this definition, the infected compartments are $A_1, I_1, I_2, C_1, C_2$, and $D$.

The Violence-Free Equilibrium (VFE) does not represent a society completely free from ideological grievances, political discontent, or latent support for violent actors. Rather, it represents a state in which no abductees undergoing radicalisation, active violent actors, captured fighters, or deradicalised former combatants are present. Thus,

$$A_1 = I_1 = I_2 = C_1 = C_2 = D = 0. \tag{12}$$

However, the sympathiser compartments $P_1$ and $P_2$ may remain active at equilibrium. Individuals in these compartments are not considered part of the violent-actor life cycle. Instead, they represent higher-risk civilian populations that are susceptible to future recruitment, radicalisation, or abduction. Individuals in $P_1$ are susceptible to voluntary radicalisation into Group 1, while individuals in both $P_1$ and $P_2$ remain vulnerable to abduction and subsequent forced radicalisation by Group 1. By contrast, individuals in the general population compartment $S$ are not directly susceptible to voluntary radicalisation until they first transition into either $P_1$ or $P_2$, although they remain vulnerable to abduction by active violent actors.

The persistence of positive equilibrium values for $P_1$ and $P_2$ arises from the baseline sympathy-generation processes $\nu_1$ and $\nu_2^{\text{eff}}$, which may continue to operate even in the absence of active violence. These processes represent the continued existence of ideological sympathy, political grievances, social discontent, or other factors that increase vulnerability to future recruitment.

Therefore, the VFE should be interpreted as a *violent-actor-free* or *violence-free* state rather than a state completely devoid of extremist, ideological, criminal, or anti-state sentiment. In practical terms, the equilibrium represents a society in which latent pools of susceptible sympathisers may persist, but no active mechanisms exist to sustain violent recruitment, forced radicalisation, mobilisation, or armed operations.

We derive the basic ('insecurity') reproduction number using the next-generation matrix approach.

Substituting equation (12) into the governing equations, the remaining non-violent compartments satisfy

$$0 = \Lambda - \nu_1 S - \nu_2^{\text{eff}} S - \mu S, \tag{13}$$

$$0 = \nu_1 S - \mu P_1, \tag{14}$$

$$0 = \nu_2^{\text{eff}} S - \mu P_2. \tag{15}$$

Solving these equations gives

$$S^0 = \frac{\Lambda}{\mu + \nu_1 + \nu_2^{\text{eff}}},$$

and

$$P_1^0 = \frac{\nu_1 S^0}{\mu}, \qquad P_2^0 = \frac{\nu_2^{\text{eff}} S^0}{\mu}.$$

Therefore, the Violence-Free Equilibrium is

$$\mathscr{E}_0 = \left(S^0,\ P_1^0,\ P_2^0,\ 0,\ 0,\ 0,\ 0,\ 0,\ 0,\ 0\right). \tag{16}$$

The total population at the VFE is

$$N_0 = S^0 + P_1^0 + P_2^0. \tag{17}$$

Under the definition adopted in this study, infection corresponds to entry into the violent-actor life cycle. The infected-state vector is

$$X = \begin{pmatrix} A_1 \\ I_1 \\ I_2 \\ C_1 \\ C_2 \\ D \end{pmatrix}. \tag{18}$$

The compartment $A_2$ represents abducted civilians who do not undergo radicalisation and do not progress directly to violent actor groups. Consequently, $A_2$ is excluded from the infected subsystem used in the next-generation matrix construction.

The infected subsystem satisfies

$$\frac{dX}{dt} = \mathscr{F}(X) - \mathscr{V}(X). \tag{19}$$

In this model, the infected subsystem is

$$\frac{dA_1}{dt} = \frac{\alpha_1 (S + P_1 + P_2) I_1}{N} - k_1 A_1, \tag{20}$$

$$\frac{dI_1}{dt} = \frac{\beta_1 P_1 I_1}{N} + \gamma_1 A_1 + \eta_1^X C_1 + \sigma_1 D - k_3 I_1, \tag{21}$$

$$\frac{dI_2}{dt} = \frac{\beta_2 P_2 I_2}{N} + \eta_2^X C_2 + \sigma_2 D - k_4 I_2, \tag{22}$$

$$\frac{dC_1}{dt} = \kappa_1 I_1 - k_5 C_1, \tag{23}$$

$$\frac{dC_2}{dt} = \kappa_2 I_2 - k_6 C_2, \tag{24}$$

$$\frac{dD}{dt} = \delta_1 C_1 + \delta_2 C_2 - k_7 D, \tag{25}$$

where

$$k_1 = \gamma_1 + \eta_1^X + \theta_1^{\text{eff}} + \mu, \tag{26}$$

$$k_3 = \kappa_1 + e_1 + \mu, \tag{27}$$

$$k_4 = \kappa_2 + e_2 + \mu, \tag{28}$$

$$k_5 = \eta_1^X + \delta_1 + \mu, \tag{29}$$

$$k_6 = \eta_2^X + \delta_2 + \mu, \tag{30}$$

$$k_7 = d + \sigma_1 + \sigma_2 + \mu. \tag{31}$$

Hence the new recruitment vector is

$$\mathscr{F}(X) = \begin{pmatrix} \dfrac{\alpha_1(S+P_1+P_2)I_1}{N} \\ \dfrac{\beta_1 P_1 I_1}{N} \\ \dfrac{\beta_2 P_2 I_2}{N} \\ 0 \\ 0 \\ 0 \end{pmatrix}. \tag{32}$$

The vector representing transition to other compartments different from new infections, $\mathscr{V}$ is given by

$$\mathscr{V}(X) = \begin{pmatrix} k_1 A_1 \\ k_3 I_1 - \gamma_1 A_1 - \eta_1^X C_1 - \sigma_1 D \\ k_4 I_2 - \eta_2^X C_2 - \sigma_2 D \\ k_5 C_1 - \kappa_1 I_1 \\ k_6 C_2 - \kappa_2 I_2 \\ k_7 D - \delta_1 C_1 - \delta_2 C_2 \end{pmatrix}. \tag{33}$$

We construct the Jacobian Matrices by taking the partial derivatives of equations (32, 33) with respect to the violent compartments in the order $A_1, I_1, I_2, C_1, C_2, D$, and evaluate at the Violence-Free Equilibrium.

At the VFE, $S^0 + P_1^0 + P_2^0 = N_0$. Therefore

$$F = D\mathscr{F}(\mathscr{E}_0) = \begin{pmatrix} 0 & \alpha_1 & 0 & 0 & 0 & 0 \\ 0 & \frac{\beta_1 P_1^0}{N_0} & 0 & 0 & 0 & 0 \\ 0 & 0 & \frac{\beta_2 P_2^0}{N_0} & 0 & 0 & 0 \\ 0 & 0 & 0 & 0 & 0 & 0 \\ 0 & 0 & 0 & 0 & 0 & 0 \\ 0 & 0 & 0 & 0 & 0 & 0 \end{pmatrix}, \tag{34}$$

and

$$V = D\mathscr{V}(\mathscr{E}_0) = \begin{pmatrix} k_1 & 0 & 0 & 0 & 0 & 0 \\ -\gamma_1 & k_3 & 0 & -\eta_1^X & 0 & -\sigma_1 \\ 0 & 0 & k_4 & 0 & -\eta_2^X & -\sigma_2 \\ 0 & -\kappa_1 & 0 & k_5 & 0 & 0 \\ 0 & 0 & -\kappa_2 & 0 & k_6 & 0 \\ 0 & 0 & 0 & -\delta_1 & -\delta_2 & k_7 \end{pmatrix}. \tag{35}$$

Because the deradicalised compartment $D$ is shared between both armed groups, and by the assumption in this model, relapse dynamics induce indirect coupling between Group 1 and Group 2 through the pathways $I_1 \to C_1 \to D \to I_2$, and $I_2 \to C_2 \to D \to I_1$.

Consequently, the next-generation matrix cannot be decomposed into two independent subsystems, and the overall basic insecurity reproduction number must be obtained from the full coupled system.

The next-generation matrix formula is given by

$$K = FV^{-1}.$$

Since the new-infection matrix $F$ contains non-zero entries only in some columns, rather than computing the full inverse explicitly, it is sufficient to determine the columns of $V^{-1}$ associated with the reproducing compartments $I_1$ and $I_2$. Thus, we require the columns 2 and 3 of $V^{-1}$ corresponding to the reproducing compartments $I_1$ and $I_2$.

***Column 2 of*** $V^{-1}$

Let

$$Vy = e_2, \tag{36}$$

where

$$e_2 = (0,1,0,0,0,0)^T, \tag{37}$$

and write

$$y = \begin{pmatrix} y_1 \\ y_2 \\ y_3 \\ y_4 \\ y_5 \\ y_6 \end{pmatrix}. \tag{38}$$

This gives the system

$$k_1 y_1 = 0, \tag{39}$$
$$-\gamma_1 y_1 + k_3 y_2 - \eta_1^X y_4 - \sigma_1 y_6 = 1, \tag{40}$$
$$k_4 y_3 - \eta_2^X y_5 - \sigma_2 y_6 = 0, \tag{41}$$
$$-\kappa_1 y_2 + k_5 y_4 = 0, \tag{42}$$
$$-\kappa_2 y_3 + k_6 y_5 = 0, \tag{43}$$
$$-\delta_1 y_4 - \delta_2 y_5 + k_7 y_6 = 0. \tag{44}$$

From equation (39),

$$y_1 = 0. \tag{45}$$

From equation (42),

$$y_4 = \frac{\kappa_1}{k_5} y_2. \tag{46}$$

From equation (43),

$$y_5 = \frac{\kappa_2}{k_6} y_3. \tag{47}$$

Substituting $y_4$ and $y_5$ into equation (44) gives

$$y_6 = \frac{\delta_1 \kappa_1}{k_5 k_7} y_2 + \frac{\delta_2 \kappa_2}{k_6 k_7} y_3. \tag{48}$$

Substituting $y_4$ and $y_6$ into equation (40) yields

$$k_3 y_2 - \frac{\eta_1^X \kappa_1}{k_5} y_2 - \frac{\sigma_1 \delta_1 \kappa_1}{k_5 k_7} y_2 - \frac{\sigma_1 \delta_2 \kappa_2}{k_6 k_7} y_3 = 1. \tag{49}$$

Hence

$$\left(k_3 - \frac{\eta_1^X \kappa_1}{k_5} - \frac{\sigma_1 \delta_1 \kappa_1}{k_5 k_7}\right) y_2 - \frac{\sigma_1 \delta_2 \kappa_2}{k_6 k_7} y_3 = 1. \tag{50}$$

Substituting $y_5$ and $y_6$ equation (41) gives

$$-\frac{\sigma_2 \delta_1 \kappa_1}{k_5 k_7} y_2 + \left(k_4 - \frac{\eta_2^X \kappa_2}{k_6} - \frac{\sigma_2 \delta_2 \kappa_2}{k_6 k_7}\right) y_3 = 0. \tag{51}$$

We define from equation (50)

$$\Delta_1 = k_3 - \frac{\eta_1^X \kappa_1}{k_5} - \frac{\sigma_1 \delta_1 \kappa_1}{k_5 k_7},$$

and from equation (51)

$$\Delta_2 = k_4 - \frac{\eta_2^X \kappa_2}{k_6} - \frac{\sigma_2 \delta_2 \kappa_2}{k_6 k_7}.$$

Then from equation (50), we have

$$\Delta_1 y_2 - \frac{\sigma_1 \delta_2 \kappa_2}{k_6 k_7} y_3 = 1,$$

And from equation (51), we have

$$-\frac{\sigma_2 \delta_1 \kappa_1}{k_5 k_7} y_2 + \Delta_2 y_3 = 0.$$

This may be written as

$$\begin{pmatrix} \Delta_1 & -\dfrac{\sigma_1 \delta_2 \kappa_2}{k_6 k_7} \\ -\dfrac{\sigma_2 \delta_1 \kappa_1}{k_5 k_7} & \Delta_2 \end{pmatrix} \begin{pmatrix} y_2 \\ y_3 \end{pmatrix} = \begin{pmatrix} 1 \\ 0 \end{pmatrix}. \tag{52}$$

The determinant of the coefficient matrix is

$$\Omega = \Delta_1 \Delta_2 - \frac{\sigma_1 \sigma_2 \delta_1 \delta_2 \kappa_1 \kappa_2}{k_5 k_6 k_7^2}. \tag{53}$$

Applying Cramer's rule gives

$$y_2 = \frac{\Delta_2}{\Omega},$$

and

$$y_3 = \frac{\sigma_2 \delta_1 \kappa_1}{k_5 k_7} \frac{1}{\Omega}.$$

Therefore,

$$(V^{-1})_{22} = \frac{\Delta_2}{\Omega}, \tag{54}$$

and

$$(V^{-1})_{32} = \frac{\sigma_2 \delta_1 \kappa_1}{k_5 k_7} \frac{1}{\Omega}. \tag{55}$$

###### 2.0.1 Column 3 of $V^{-1}$

Let

$$Vz = e_3, \tag{56}$$

where

$$e_3 = (0,0,1,0,0,0)^T. \tag{57}$$

Proceeding exactly as above gives

$$(V^{-1})_{23} = \frac{\sigma_1 \delta_2 \kappa_2}{k_6 k_7} \frac{1}{\Omega}, \tag{58}$$

and

$$(V^{-1})_{33} = \frac{\Delta_1}{\Omega}. \tag{59}$$

Hence the $I_1$-$I_2$ block of $V^{-1}$ is

$$\begin{pmatrix} (V^{-1})_{22} & (V^{-1})_{23} \\ (V^{-1})_{32} & (V^{-1})_{33} \end{pmatrix} = \frac{1}{\Omega} \begin{pmatrix} \Delta_2 & \dfrac{\sigma_1 \delta_2 \kappa_2}{k_6 k_7} \\ \dfrac{\sigma_2 \delta_1 \kappa_1}{k_5 k_7} & \Delta_1 \end{pmatrix}. \tag{60}$$

Because only $I_1$ and $I_2$ appear in non-zero columns of VFE, the spectral radius of $FV^1$ equals that of the reduced 2×2 Schur complement.

The reduced next-generation matrix is therefore

$$M = \begin{pmatrix} \dfrac{\beta_1 P_1^0}{N_0} + \dfrac{\alpha_1 \gamma_1}{k_1} & 0 \\ 0 & \dfrac{\beta_2 P_2^0}{N_0} \end{pmatrix} \frac{1}{\Omega} \begin{pmatrix} \Delta_2 & \dfrac{\sigma_1 \delta_2 \kappa_2}{k_6 k_7} \\ \dfrac{\sigma_2 \delta_1 \kappa_1}{k_5 k_7} & \Delta_1 \end{pmatrix}. \tag{61}$$

Multiplying gives

$$M = \frac{1}{\Omega} \begin{pmatrix} \left(\frac{\beta_1 P_1^0}{N_0} + \frac{\alpha_1 \gamma_1}{k_1}\right) \Delta_2 & \left(\frac{\beta_1 P_1^0}{N_0} + \frac{\alpha_1 \gamma_1}{k_1}\right) \dfrac{\sigma_1 \delta_2 \kappa_2}{k_6 k_7} \\ \frac{\beta_2 P_2^0}{N_0} \dfrac{\sigma_2 \delta_1 \kappa_1}{k_5 k_7} & \frac{\beta_2 P_2^0}{N_0} \Delta_1 \end{pmatrix}. \tag{62}$$

Define

$$R_{11} = \frac{\left(\frac{\beta_1 P_1^0}{N_0} + \frac{\alpha_1 \gamma_1}{k_1}\right) \Delta_2}{\Omega},$$

$$R_{12} = \frac{\left(\frac{\beta_1 P_1^0}{N_0} + \frac{\alpha_1 \gamma_1}{k_1}\right) \dfrac{\sigma_1 \delta_2 \kappa_2}{k_6 k_7}}{\Omega},$$

$$R_{21} = \frac{\frac{\beta_2 P_2^0}{N_0} \frac{\sigma_2 \delta_1 \kappa_1}{k_5 k_7}}{\Omega},$$

$$R_{22} = \frac{\frac{\beta_2 P_2^0}{N_0} \Delta_1}{\Omega}.$$

Thus,

$$M = \begin{pmatrix} R_{11} & R_{12} \\ R_{21} & R_{22} \end{pmatrix}, \tag{63}$$

and

$$R_0 = \rho(M),$$

where $\rho(M)$ denotes the spectral radius of the next-generation matrix. This gives

$$R_0 = \frac{R_{11} + R_{22} + \sqrt{(R_{11} - R_{22})^2 + 4R_{12}R_{21}}}{2}. \tag{64}$$

#### 3 Humanitarian (Civilian) Harm Index

To evaluate the humanitarian consequences of violent insecurity, we introduce an instantaneous civilian harm function that quantifies the overall burden imposed on civilians at time $t$. This is an 'index' in the sense that it is a single summary statistic derived from weighting various measures at time $t$, whose changes can be tracked over time, and which can be used as a policy target. The formulation is designed to capture the real-time intensity of humanitarian loss and therefore provides a metric for evaluating the effectiveness of time-dependent intervention strategies.

In this study, humanitarian or civilian harm is associated with realised adverse outcomes experienced by civilians rather than the duration of captivity itself. Consequently, time spent in captivity is not treated as an independent source of humanitarian burden. Instead, captivity contributes indirectly through observable events and outcomes experienced by abducted civilians.

Accordingly, the model recognises four complementary dimensions of humanitarian harm:

1. civilian mortality resulting from violence and captivity;
2. new civilian abductions;
3. coerced radicalisation of abducted civilians;
4. ransom extraction from abducted civilians.

These dimensions represent distinct forms of humanitarian loss and need not contribute equally to overall societal burden. We therefore introduce positive weighting coefficients satisfying

$$w_D > 0, \qquad w_A > 0, \qquad w_R > 0, \qquad w_Q > 0, \tag{65}$$

where

- $w_D$: mortality-related humanitarian burden,
- $w_A$: abduction-related humanitarian burden,
- $w_R$: forced radicalisation burden,
- $w_Q$: ransom-related humanitarian burden.

##### 3.1 Civilian Mortality Rate

We define the instantaneous civilian mortality rate as

$$M(t) = m_1 \frac{(S+P_1+P_2)I_1}{N} + m_2 \frac{(S+P_1+P_2)I_2}{N} + \theta_1^{\text{eff}} A_1 + \theta_2^{\text{eff}} A_2. \tag{66}$$

The first two terms represent direct civilian deaths generated by violent attacks carried out by active members of Groups 1 and 2. The final two terms represent deaths occurring among abducted civilians while in captivity.

Thus, $M(t)$ measures the total instantaneous rate of civilian mortality generated by both direct violence and captivity-related deaths.

##### 3.2 Abduction Incidence Rate

We define the instantaneous civilian abduction rate as

$$A(t) = \alpha_1 \frac{(S+P_1+P_2)I_1}{N} + \alpha_2 \frac{(S+P_1+P_2)I_2}{N}. \tag{67}$$

This quantity measures the rate at which civilians are newly abducted by the two armed groups and therefore captures the immediate kidnapping risk faced by the civilian population.

Abduction is included directly because kidnapping itself constitutes an immediate humanitarian event regardless of subsequent outcomes.

##### 3.3 Forced Radicalisation Burden

We recognise that some abducted civilians may be transformed into active violent participation through coercion. We therefore define the forced radicalisation burden as

$$R_H(t) = \gamma_1 A_1. \tag{68}$$

This quantity measures the instantaneous rate at which abducted civilians held by Group 1 are converted into active members of the group against their will.

##### 3.4 Ransom Burden

The model further recognises ransom extraction as a distinct humanitarian burden.

We therefore define the ransom burden as

$$Q_H(t) = \eta_2^R A_2. \tag{69}$$

This quantity measures the instantaneous rate at which abducted civilians held by Group 2 generate ransom incentives.

Although ransom may eventually lead to civilian release, it may impose substantial humanitarian costs through economic deprivation, household financial distress, uncertainty, coercion, disruption of livelihoods, and broader social instability. Accordingly, ransom extraction is treated as an independent humanitarian outcome.

##### 3.5 Weighted Humanitarian Harm

Combining the four dimensions of humanitarian impact, we define the overall instantaneous humanitarian harm function by

$$h(t) = w_D M(t) + w_A A(t) + w_R R_H(t) + w_Q Q_H(t). \tag{70}$$

Substituting the component definitions yields

$$\begin{aligned} h(t) = w_D &\left[ m_1 \frac{(S+P_1+P_2)I_1}{N} + m_2 \frac{(S+P_1+P_2)I_2}{N} + \theta_1^{\text{eff}} A_1 + \theta_2^{\text{eff}} A_2 \right] \\ &+ w_A \left[ \alpha_1 \frac{(S+P_1+P_2)I_1}{N} + \alpha_2 \frac{(S+P_1+P_2)I_2}{N} \right] \\ &+ w_R \gamma_1 A_1 + w_Q \eta_2^R A_2. \end{aligned} \tag{71}$$

This formulation combines mortality, abduction activity, forced radicalisation, and ransom extraction into a single policy-relevant indicator while avoiding direct dependence on captivity duration.

##### 3.6 Normalised Humanitarian Harm

To account for temporal variation in population size, we define the civilian population as

$$N_c(t) = S(t) + P_1(t) + P_2(t) + A_1(t) + A_2(t). \tag{72}$$

The corresponding normalised humanitarian harm rate is defined by

$$h_c(t) = \frac{h(t)}{N_c(t)}. \tag{73}$$

In this study, we refer to $h_c(t)$ as the Humanitarian Harm Index or *Civilian Harm Index (CHI)*. This quantity represents the per-capita humanitarian burden experienced by civilians per unit time.

##### 3.7 Cumulative Humanitarian Burden

For sensitivity analysis and policy evaluation, we also quantified the cumulative humanitarian burden over the simulation time $[0, T]$ using

$$H(T) = \int_0^T h_c(t)\, \mathrm{d}t. \tag{74}$$

We refer to $H(T)$ as the *Cumulative Harm Burden* or (in the main article) the *cumulative Civilian Harm Index*. This quantity measures the cumulative per-capita humanitarian burden generated throughout the conflict period (time 0–T) and provides a metric for comparing alternative intervention strategies on reducing this quantity.

Under successful intervention policies, the normalised humanitarian harm rate is expected to decline over time as violence, kidnapping activity, captivity-related mortality, forced recruitment, and ransom-driven exploitation are suppressed.

#### 4 Global Sensitivity Analysis of the basic insecurity Reproduction Number and Harm Index

To evaluate parameter uncertainty and identify the dominant mechanisms driving violent insecurity outcomes, a Global Sensitivity Analysis (GSA) was conducted on two principal model outputs:

1. the basic insecurity reproduction number, $R_0$, which quantifies the transmission potential and persistence of violent activity; and
2. the cumulative Harm Index, $H$, which measures the aggregate humanitarian burden generated by violence.

The analysis was performed on the full frequency-dependent multi-actor ODE system across selected policy-relevant parameters.

A total of 17 parameters were included:

$$\beta_1,\ \beta_2,\ \nu_1,\ \nu_2,\ \alpha_1,\ \alpha_2,\ \gamma_1,\ \kappa_1,\ \kappa_2,\ e_1,\ e_2,\ \delta_1,\ \delta_2,\ \sigma_1,\ \eta_1^X,\ \eta_2^R,\ \psi$$

where recruitment, radicalisation, intervention effectiveness, mortality, and ransom-related mechanisms were all treated as uncertain inputs.

Each parameter was sampled independently over a uniform interval corresponding to $\pm 50\%$ of its baseline value:

$$\theta_i \sim U(0.5\theta_i^{(0)}, 1.5\theta_i^{(0)}) \tag{75}$$

where $\theta_i^{(0)}$ denotes the baseline estimate.

Sampling was performed using Latin Hypercube Sampling (LHS) with 500 parameter sets for each parameter to ensure efficient exploration of the multidimensional parameter space while maintaining computational tractability.

###### *4.0.1 Sensitivity Analysis of* $R_0$

For each sampled parameter set, the basic insecurity reproduction number was recomputed using the exact next-generation formulation in equation (63).

This procedure generated an empirical distribution of $R_0$ values across the uncertainty space.

##### 4.1 Sensitivity Analysis of the Harm Index

The (CHI) was analysed separately because it represents. For each sampled parameter set, the full ODE system was simulated over the study time using the `lsoda` numerical solver. The response variable was defined as the cumulative CHI, and was then evaluated for the total time $(T)$ as using equation (74), which was approximated numerically using trapezoidal integration over discrete simulation time points.

Global sensitivity of $H(T)$ was quantified using Spearman rank correlations. For Spearman analysis, negative coefficients indicate that increasing the parameter of interest's values are associated with reduced cumulative CHI, whereas positive coefficients indicate increased CHI.

##### 4.2 Intervention implementation and evaluation

Intervention scenarios were implemented by modifying selected model parameters that represent alternative policy responses and re-running the full ODE system. Each intervention acts directly on one or more behavioural or operational mechanisms in the model rather than imposing an external control term.

Specifically, interventions were represented as parameter adjustments of $\pm 50\%$ of the baseline values. Parameters whose reduction represent preventive actions $(v_1, v_2)$ were reduced by 50%, whereas parameters representing intervention intensity; prisoner exchange rates $(\eta_1^X, \eta_2^X)$, ransom-mediated release $(\eta_2^R)$, arrest intensity $(\kappa_1, \kappa_2)$, de-radicalisation $(\delta_1, \delta_2)$, and military elimination of active violent members $(e_1, e_2)$ were increased by 50%.. For each scenario, the modified parameter set was passed into the model solver while all other parameters and initial conditions remained unchanged.

Intervention effectiveness was evaluated using the humanitarian harm function rather than epidemiological prevalence alone. At each simulation time step, an instantaneous per-capita civilian harm measure.

To assess intervention effectiveness consistently across simulations, the percentage change in Harm Index relative to the baseline scenario was computed as:

$$\Delta H = 100\left(1 - \frac{H_{\text{scenario}}}{H_{\text{baseline}}}\right), \tag{76}$$

where $H_{\text{baseline}}$ is the harm index under the baseline conditions and $H_{\text{scenario}}$ is the harm index under the intervention scenario. Positive values of $\Delta H$ indicate a reduction in civilian harm relative to baseline, whereas negative values indicate increased harm/burden.

##### 4.3 Bifurcation Analysis

The basic insecurity reproduction number $R_0$ determines the threshold governing invasion and persistence of violent insecurity. While the next-generation matrix establishes local stability of the Violence-Free Equilibrium, it does not determine the qualitative dynamics of the system near the critical threshold $R_0 = 1$. To characterise local behaviour near this threshold, we apply centre manifold theory following Castillo-Chavez and Song.

Let

$$X = (S, P_1, P_2, A_1, A_2, I_1, I_2, C_1, C_2, D)^T$$

denote the state vector and write the system compactly as

$$\frac{dX}{dt} = F(X, \beta_1), \tag{77}$$

where $\beta_1$ is selected as the bifurcation parameter.

Let $\beta_1 = \beta_1^*$ denote the critical parameter value satisfying

$$R_0(\beta_1^*) = 1. \tag{78}$$

At this threshold, assume that the Jacobian matrix

$$J(A_0, \beta_1^*)\ \frac{\partial F}{\partial X}\bigg|_{(A_0, \beta_1^*)} \tag{79}$$

possesses a simple zero eigenvalue while all remaining eigenvalues have negative real parts.

Let

$$w = (w_1, \ldots, w_{10})^T$$

and

$$v = (v_1, \dots, v_{10})$$

denote the associated right and left eigenvectors satisfying

$$J(A_0, \beta_1^*)w = 0,$$

and

$$v^T J(A_0, \beta_1^*) = 0,$$

with normalisation

$$v \cdot w = 1.$$

Following Castillo-Chavez and Song, define

$$a = \sum_{k=1}^{10} \sum_{i=1}^{10} \sum_{j=1}^{10} v_k w_i w_j \frac{\partial^2 F_k}{\partial x_i \partial x_j}(A_0, \beta_1^*), \tag{80}$$

and

$$b = \sum_{k=1}^{10} \sum_{i=1}^{10} v_k w_i \frac{\partial^2 F_k}{\partial x_i \partial \beta_1}(A_0, \beta_1^*). \tag{81}$$

Only nonlinear recruitment and interaction terms contribute to these second derivatives.

Since $\beta_1$ appears only in the recruitment term of the active Group 1 equation,

$$\frac{dI_1}{dt} = \beta_1 \frac{P_1 I_1}{N} + \gamma_1 A_1 + \eta_1^X C_1 + \sigma_1 D - (\kappa_1 + e_1 + \mu) I_1, \tag{82}$$

it follows that

$$\frac{\partial^2 F_6}{\partial I_1 \partial \beta_1} = \frac{P_1}{N}. \tag{83}$$

Evaluating at the VFE gives

$$\left.\frac{\partial^2 F_6}{\partial I_1 \partial \beta_1}\right|_{A_0} = \frac{P_1^0}{N^0}. \tag{84}$$

Therefore,

$$b = v_6 w_6 \frac{P_1^0}{N^0}. \tag{85}$$

Since

$$v_6 > 0, \qquad w_6 > 0, \qquad P_1^0 > 0, \qquad N^0 > 0,$$

we obtain

$$b > 0.$$

The direction of bifurcation therefore depends entirely on the sign of the first centre manifold coefficient $a$. If

$$a < 0,$$

the system undergoes a forward (supercritical) transcritical bifurcation and the VFE remains locally asymptotically stable for $R_0 < 1$, while a stable endemic equilibrium emerges for $R_0 > 1$. If

$$a > 0,$$

the system exhibits backward bifurcation, allowing coexistence of violence-free and endemic equilibria for some parameter regimes satisfying $R_0 < 1$.

Since the relapse mechanism introduces coupling between the two armed groups through the pathway

$$I_g \to C_g \to D \to I_h, \qquad g,h \in \{1,2\},$$

the associated centre eigenvectors depend non-trivially on the parameters $(\sigma_1, \sigma_2, \delta_1, \delta_2, \kappa_1, \kappa_2)$. Although relapse contributes linearly to the governing equations and therefore does not generate direct Hessian terms, it modifies the centre manifold coefficient through the associated eigenvector structure.

Consequently, the sign of $a$ cannot be determined directly from structural arguments alone. Therefore, this analysis establishes that

$$b > 0,$$

while the direction of bifurcation remains governed by the sign of $a$.

Explicit evaluation of $a$ would require symbolic computation of the Hessian tensor together with numerical construction of the associated left and right eigenvectors at the critical parameter value.

##### 4.4 Single-Parameter Threshold Exploration

This framework models two interacting violent groups operating through partially independent recruitment and persistence pathways. This would mean that reducing the overall reproduction potential below unity may not be achievable through isolated intervention mechanisms (for example, targeting $\beta_1$ used as the bifurcation parameter).

To investigate this numerically, a systematic single-parameter threshold exploration was performed. Each controllable parameter was varied independently while all remaining parameters were fixed at their baseline values. Parameters representing recruitment, sympathy generation, contact, and relapse were reduced from their baseline values to their theoretical upper bound of zero, whereas control-related parameters (capture, elimination, deradicalisation, reintegration, negotiation, and captivity exit) were increased from baseline values up to their theoretical upper bound of one. These lower and upper bounds are regimes where these parameters have the highest effect on reducing the $R_0$.

For each intervention scenario, the basic insecurity reproduction number $R_0$ was recomputed and the minimum achievable value recorded. The objective was to determine whether any individual intervention could independently force the system below the elimination threshold $(R_0 < 1)$.

Results are shown in Table 2. Across all parameters explored, no single intervention succeeded in reducing $R_0$ below unity. The lowest value obtained was $R_0 = 1.118$, indicating that violent persistence in the present multi-actor system is sustained through multiple concurrent transmission and recruitment mechanisms. This observation suggests that threshold reduction requires coordinated intervention across several mechanisms simultaneously rather than isolated control actions.

The numerical exploration showed that no single intervention parameter was sufficient to drive $R_0$ below unity. This suggests that the bifurcation point is not accessible through isolated control actions under the baseline regime. Instead, persistence is maintained by the coupled multi-actor structure of the system, implying that coordinated multi-parameter intervention strategies are required to reach the elimination threshold.

##### 4.5 Numerical Simulation and Baseline Parameterisation

To investigate the behaviour of the proposed violent insecurity model and evaluate humanitarian outcomes, numerical simulations were performed in `R` using the `deSolve` package to run system of ordinary differential equations. The ODEs was solved using the adaptive `lsoda` algorithm, which automatically switches between stiff and non-stiff integration methods depending on the local characteristics of the system. Numerical integration was conducted over the interval

$$t \in [0, 1500],$$

using a step increment of 0.2 time units and solver tolerances

$$\texttt{rtol} = 10^{-8}, \qquad \texttt{atol} = 10^{-10},$$

with a maximum of 500000 integration steps to ensure numerical stability and convergence.

**Table 2.** Single-parameter intervention screening for threshold reduction. Each parameter was varied independently while all other parameters remained fixed at baseline values. Reduction parameters were explored from baseline to (0), while enhancement parameters were explored from baseline to (1).

| Parameter | Baseline | Search Range | Minimum $(R_0)$ |
|---|---|---|---|
| $\alpha_1$ | 0.060 | $[0, 0.060]$ | 1.861 |
| $\alpha_2$ | 0.050 | $[0, 0.050]$ | 2.496 |
| $\beta_1$ | 0.230 | $[0, 0.230]$ | 1.753 |
| $\beta_2$ | 0.250 | $[0, 0.250]$ | 2.487 |
| $d$ | 0.080 | $[0.080, 1]$ | 2.378 |
| $\delta_1$ | 0.060 | $[0.060, 1]$ | 1.773 |
| $\delta_2$ | 0.050 | $[0.050, 1]$ | 2.491 |
| $e_1$ | 0.015 | $[0.015, 1]$ | 1.749 |
| $e_2$ | 0.012 | $[0.012, 1]$ | 2.483 |
| $\eta_{1X}$ | 0.060 | $[0.060, 1]$ | 2.496 |
| $\eta_{2X}$ | 0.050 | $[0.050, 1]$ | 2.496 |
| $\eta_{2R}$ | 0.080 | $[0.080, 1]$ | 2.346 |
| $\kappa_1$ | 0.080 | $[0.080, 1]$ | 1.753 |
| $\kappa_2$ | 0.060 | $[0.060, 1]$ | 2.489 |
| $\nu_1$ | 0.0015 | $[0, 0.0015]$ | 2.136 |
| $\nu_2$ | 0.0010 | $[0, 0.0010]$ | 2.496 |
| $\sigma_1$ | 0.006 | $[0, 0.006]$ | 2.368 |
| $\sigma_2$ | 0.004 | $[0, 0.004]$ | 2.489 |
| $\theta_1$ | 0.020 | $[0.020, 1]$ | 1.958 |
| $\theta_2$ | 0.010 | $[0.010, 1]$ | 2.496 |

The initial population state was specified as

$$(S(0), P_1(0), P_2(0), A_1(0), A_2(0), I_1(0), I_2(0), C_1(0), C_2(0), D(0)) = (500000, 20000, 15000, 0, 0, 50, 40, 0, 0, 0).$$

This configuration represents a predominantly susceptible population with existing ideological support populations and a small initial active insecurity presence.

Baseline parameter values are summarised in Table 3.

##### 4.6 Numerical Computation and Stability of the Violence Persistence Equilibrium

To characterise the long-term behaviour of the insecurity system beyond threshold analysis, the violence persistence equilibrium (VPE) was computed numerically by setting all model derivatives equal to zero and solving the resulting steady-state system $(F(X) = 0)$. The resulting nonlinear algebraic system was solved simultaneously using a multidimensional root-finding procedure implemented through the `multiroot()` routine in `R`. Since the resulting equations could not be solved analytically, a numerical root-finding procedure was used to search for values of all state variables that make all equations simultaneously equal to zero. To assess the existence of multiple steady states, the computation was repeated from multiple initial conditions. The numerical search consistently converged to a single feasible positive equilibrium in addition to the analytically derived Violence-Free Equilibrium. The equilibrium solution converged with negligible residual error ($< 10^{-12}$), confirming numerical consistency of the computed steady state.

The resulting violence persistence equilibrium is

$$E^* = (S^*, P_1^*, P_2^*, A_1^*, A_2^*, I_1^*, I_2^*, C_1^*, C_2^*, D^*) = \begin{pmatrix} 401776.20 \\ 59487.10 \\ 93785.99 \\ 6596.87 \\ 2727.65 \\ 25169.18 \\ 8762.48 \\ 16640.78 \\ 5205.44 \\ 13832.07 \end{pmatrix}. \quad (86)$$

**Table 3.** Model parameters, baseline values and interpretations.

| Parameter | Meaning | Value | Units |
|---|---|---|---|
| $\Lambda$ | Recruitment into the susceptible population | 800 | persons/time |
| $\mu$ | Natural mortality rate | 0.001 | 1/time |
| $\nu_1$ | Rate at which susceptibles become sympathisers of Group 1 | 0.0015 | 1/time |
| $\nu_2$ | Rate at which susceptibles become sympathisers of Group 2 | 0.0010 | 1/time |
| $\beta_1$ | Recruitment rate from sympathisers $P_1$ into active violent actors $I_1$ | 0.23 | 1/time |
| $\beta_2$ | Recruitment rate from sympathisers $P_2$ into active violent actors $I_2$ | 0.25 | 1/time |
| $\alpha_1$ | Abduction/contact rate associated with Group 1 | 0.06 | 1/time |
| $\alpha_2$ | Abduction/contact rate associated with Group 2 | 0.05 | 1/time |
| $\gamma_1$ | Radicalisation rate of abducted individuals into Group 1 | 0.12 | 1/time |
| $\eta_1^X$ | Prisoner exchange/release rate for Group 1 | 0.06 | 1/time |
| $\eta_2^X$ | Prisoner exchange/release rate for Group 2 | 0.05 | 1/time |
| $\eta_2^R$ | Ransom-release rate for Group 2 | 0.08 | 1/time |
| $\xi_1$ | Negotiation effectiveness coefficient for Group 1 | 0.50 | dimensionless |
| $\xi_2$ | Negotiation effectiveness coefficient for Group 2 | 0.30 | dimensionless |
| $\theta_1$ | Captivity exit/mortality rate among abductees of Group 1 | 0.02 | 1/time |
| $\theta_2$ | Captivity exit/mortality rate among abductees of Group 2 | 0.01 | 1/time |
| $\kappa_1$ | Capture/arrest rate of active Group 1 members | 0.08 | 1/time |
| $\kappa_2$ | Capture/arrest rate of active Group 2 members | 0.06 | 1/time |
| $e_1$ | Elimination rate of active Group 1 members | 0.015 | 1/time |
| $e_2$ | Elimination rate of active Group 2 members | 0.012 | 1/time |
| $\delta_1$ | Deradicalisation rate of captured Group 1 members | 0.06 | 1/time |
| $\delta_2$ | Deradicalisation rate of captured Group 2 members | 0.05 | 1/time |
| $d$ | Reintegration rate of recovered individuals | 0.080 | 1/time |
| $\sigma_1$ | Relapse rate into Group 1 | 0.006 | 1/time |
| $\sigma_2$ | Relapse rate into Group 2 | 0.004 | 1/time |
| $\psi$ | Recruitment amplification induced by successful ransom outcomes | 0.50 | dimensionless |
| $r$ | Population growth rate (simulation only) | 0.046 | 1/time |
| $w_D$ | Humanitarian burden weight for civilian mortality | 1.00 | dimensionless |
| $w_A$ | Humanitarian burden weight for civilian abduction | 0.30 | dimensionless |
| $w_R$ | Humanitarian burden weight for forced radicalisation | 0.50 | dimensionless |
| $w_Q$ | Humanitarian burden weight for ransom-related burden | 0.40 | dimensionless |

Since each unit is a person, values in the main text are quoted as integers rather than either quoting the fractional part or limiting the number of significant figures.

All compartments remained strictly positive at equilibrium, indicating coexistence of susceptible individuals, ideological support populations, abducted individuals, active violent actors, captured populations, and rehabilitated individuals under sustained insecurity conditions. In particular, both active insecurity compartments remained positive ($I_1^* = 25169.18$, $I_2^* = 8762.48$), demonstrating that the system converges to a persistent violence regime rather than returning to the Violence-Free Equilibrium.

Local stability of the persistence equilibrium was investigated by evaluating the Jacobian matrix at $E^*$ and computing its eigenvalues numerically. The eigenvalue spectrum obtained is

$$\lambda = \{-0.2474,\ -0.1466,\ -0.1395,\ -0.1296,\ -0.0975,\ -0.0067 \pm 0.0109i,\ -0.0055 \pm 0.0097i,\ -0.0014\}. \tag{87}$$

All eigenvalues possessed strictly negative real parts, with the dominant eigenvalue

$$\lambda_{\max} = -0.00135. \tag{88}$$

Therefore, the violence persistence equilibrium is locally asymptotically stable.
